# Functional variants in the TAS2R38 bitter taste receptor associate with postprandial glycemia

**DOI:** 10.1101/2025.05.23.25328232

**Authors:** Julie E. Gervis, Kenneth E. Westerman, Joanne B. Cole, Jordi Merino, Sara J. Cromer, Miriam S. Udler

## Abstract

**Aims/Hypothesis:** Functional variants in the TAS2R38 bitter taste receptor influence bitter taste perception and inform dietary and lifestyle behaviors that impact glucose homeostasis. Experimental data suggest that TAS2R38 receptors also mediate postprandial GLP-1 secretions from intestinal L-cells, though human study data are limited. To further establish the role of TAS2R38 in glucose homeostasis in humans, we tested whether functional variants conferring greater TAS2R38 sensitivity associate with lower blood glucose, particularly in the postprandial state, independent of dietary- and lifestyle-mediated effects of TAS2R38 on glucose levels.

**Methods:** We analyzed participants without type 2 diabetes of European ancestry in the UK Biobank. We used known functional variants in *TAS2R38* to assign canonical haplotypes (AVI, PAV) and diplotypes conferring low (AVI/AVI nontasters), moderate (AVI/PAV tasters) or high (PAV/PAV supertasters) TAS2R38 receptor sensitivity and bitter taste perception. Linear models were used to quantify the associations of TAS2R38 diplotype with random glucose, and glucose levels over various time windows spanning postprandial and fasting states, adjusting for demographics and BMI, then sequentially for dietary and lifestyle factors. We used variants in other bitter taste receptors (TAS2R14 and TAS2R19), related to similar dietary and lifestyle behaviors as TAS2R38 but without hypothesized roles in glucose metabolism, to serve as negative controls.

**Results:** Among 218,688 individuals, 34%, 49% and 18% were AVI/AVI nontasters, AVI/PAV tasters and PAV/PAV supertasters, respectively. In BMI-adjusted models, each additional PAV haplotype associated with decreases in random glucose levels (beta [95% CI] = −0.07 [−0.13, −0.01] mg/dL; *P* = 0.021). In analyses stratified by fasting time, associations were only significant in the 0-2 hr ‘postprandial’ window (−0.24 [−0.39, −0.08] mg/dL per PAV haplotype; *P* = 0.003). These associations replicated (at the variant-level) in a published GWAS meta-analysis of 2-hr OGTT glucose and persisted after further adjustment for dietary and lifestyle behaviors (adjusted beta = −0.22; *P* = 0.004). Variants in *TAS2R14* and *TAS2R19* were related to similar behavioral traits as *TAS2R38* but were not associated with 0-2 hr glucose, supporting behaviorally-independent effects of TAS2R38 diplotypes on 0-2 hr glucose.

**Conclusions/interpretation:** Functional variants conferring greater TAS2R38 receptor sensitivity were associated with lower glucose levels in the postprandial state. These findings align with experimental evidence supporting a functional role for TAS2R38 in postprandial glycemia, reinforcing it as a potential target for type 2 diabetes prevention and treatment.

**Research in Context:** *What is already known about this subject?:*

- TAS2R38 is a specialized G-protein coupled receptor that mediates bitter taste perception in the mouth and has been discovered to act as a peripheral nutrient sensor in intestinal L-cells in the gut.
- Functional variants in *TAS2R38* conferring its sensitivity give rise to three canonical diplotypes with well-defined effects on bitter taste perception and dietary and lifestyle behaviors relevant to glucose homeostasis.
- Experimental studies show that TAS2R38 receptors in human enteroendocrine L-cells contribute to the postprandial secretion of incretin hormone, glucagon-like peptide-1 (GLP-1).

*What is the key question?:*

- Are functional variants in the *TAS2R38* bitter taste receptor associated with glucose homeostasis in humans, particularly in the postprandial state?

*What are the new findings?:*

- In a large cohort of adults without type 2 diabetes, individuals carrying two copies of the haplotype encoding more sensitive TAS2R38 receptors (PAV) had significantly lower postprandial glucose levels than those with two copies of the non-functional haplotype (AVI), regardless of dietary or lifestyle behaviors, with differences following a dose-response relationship per PAV haplotype.

*How might this impact on clinical practice in the foreseeable future?:*

- These findings provide evidence to support direct TAS2R38 actions in postprandial glycemia, which could inform subsequent experimental work to leverage TAS2R38 as a therapeutic target for impaired glucose regulation.

## Introduction

A better understanding of the mechanisms contributing to glucose homeostasis is critical to identify new targets for the prevention and treatment of type 2 diabetes. Given the clinical success of glucagon-like peptide-1 (GLP-1) receptor agonist medications for improving glycemic control,^1^ gut nutrient sensing mechanisms that modulate endogenous GLP-1 secretion have become key areas of interest.^2^ Type 2 taste receptors––a specialized family of G-protein coupled receptors originally known for their role in bitter taste perception^3^––have been recently discovered in gut enteroendocrine L-cells which store and secrete GLP-1.^4–6^ Activation of one subtype, TAS2R38, by bitter-tasting ligands has been shown to trigger GLP-1 secretion in animal and human cell models in a dose-dependent manner.^7–11^ Consistent with this mechanism, in preclinical studies, several bitter-tasting ligands have been shown to promote GLP-1 and insulin secretions and reduce postprandial glucose responses to mixed-nutrient challenges in healthy adults and those with type 2 diabetes,^12–14^ supporting TAS2R38 as a target to promote endogenous GLP-1 secretion and improve glucose regulation.^8,15^

Common missense variants within the *TAS2R38* gene give rise to two canonical haplotypes with well-defined functional and behavioral implications: the *AVI* haplotype encodes for a non-functional receptor, while the *PAV* haplotype encodes for a functional receptor.^16,17^ Collectively, these variants explain ~70% of phenotypic variability in bitter taste perception by modulating the ligand-binding capacity of the receptor.^18,19^ When assessed as a diplotype, individuals with one or two copies of the functional *PAV* haplotype who have normal or heightened bitter taste perception (i.e., *AVI*/*PAV* or *PAV*/*PAV*; so called “tasters” or “supertasters,” respectively)^17^ also tend to consume less coffee, alcohol, and bitter vegetables and smoke less than those with two copies of the non-functional *AVI* haplotype who have blunted bitter taste perception (i.e., *AVI/AVI*; so called “nontasters”).^20–23^

Despite evidence implicating TAS2R38 in glucose homeostasis, only one prior study has examined the impact of functional variations in *TAS2R38* on glucose metabolism.^23^ Among ~1000 German adults without type 2 diabetes in the Sorbs cohort, genetic tasters and supertasters had significantly lower glucose at 30-min and 120-min area under the curve (AUC) after a 75 g oral glucose tolerance test (OGTT) than nontasters, despite similar fasting plasma glucose, consistent with proposed TAS2R38 roles in GLP-1 secretion. Whether these differences in postprandial glucose responses were informed by dietary or lifestyle (i.e., behavioral) effects of *TAS2R38* or whether they reflect direct biological actions of TAS2R38 in glucose homeostasis is unknown. Delineating these effects using large-scale human studies can help understand the mechanisms through *TAS2R38* contributes to glucose homeostasis and establish the clinical relevance of functional variants in TAS2R38 to type 2 diabetes prevention and treatment.

Here, we examined the associations of functional variants in the *TAS2R38* bitter taste receptor gene with markers of glucose homeostasis among adults without type 2 diabetes in a large, population-based biobank. We hypothesized that functional variants and canonical diplotypes conferring more sensitive TAS2R38 bitter taste receptors would associate with lower blood glucose levels, particularly in the postprandial state; and that these associations would be independent of dietary and lifestyle behaviors, supporting evidence of direct biological actions of TAS2R38 in postprandial glucose metabolism.

## Methods

### UK Biobank participants

The UK Biobank (UKB) is a prospective cohort which enrolled adults aged 40-69 years living throughout the UK, with comprehensive genetic, phenotypic, and health records data.^24,25^ We retrieved information on genetic relatedness, ancestry group, and principal components (PCs) from the Pan-UKB project.^26^ We excluded participants who withdrew their consent from the biobank by the time of the analysis, and included unrelated individuals of European genetic ancestry with a hemoglobin A1c (HbA1c) <5.7% (39 mmol/mol) and who did not have type 2 diabetes as defined by a published algorithm^27^ to ensure a focus on those without dysglycemia (*N* = 362,458). From this subsample, we included individuals with complete data on genotyping, glucose outcomes, dietary habits (from UKB food frequency questionnaires [FFQ]), and relevant dietary and lifestyle confounders and we removed those with extreme values (>5 SD) for glucose outcomes (see flow diagram in electronic supplementary material [ESM] Fig. 1).

**Figure 1.**
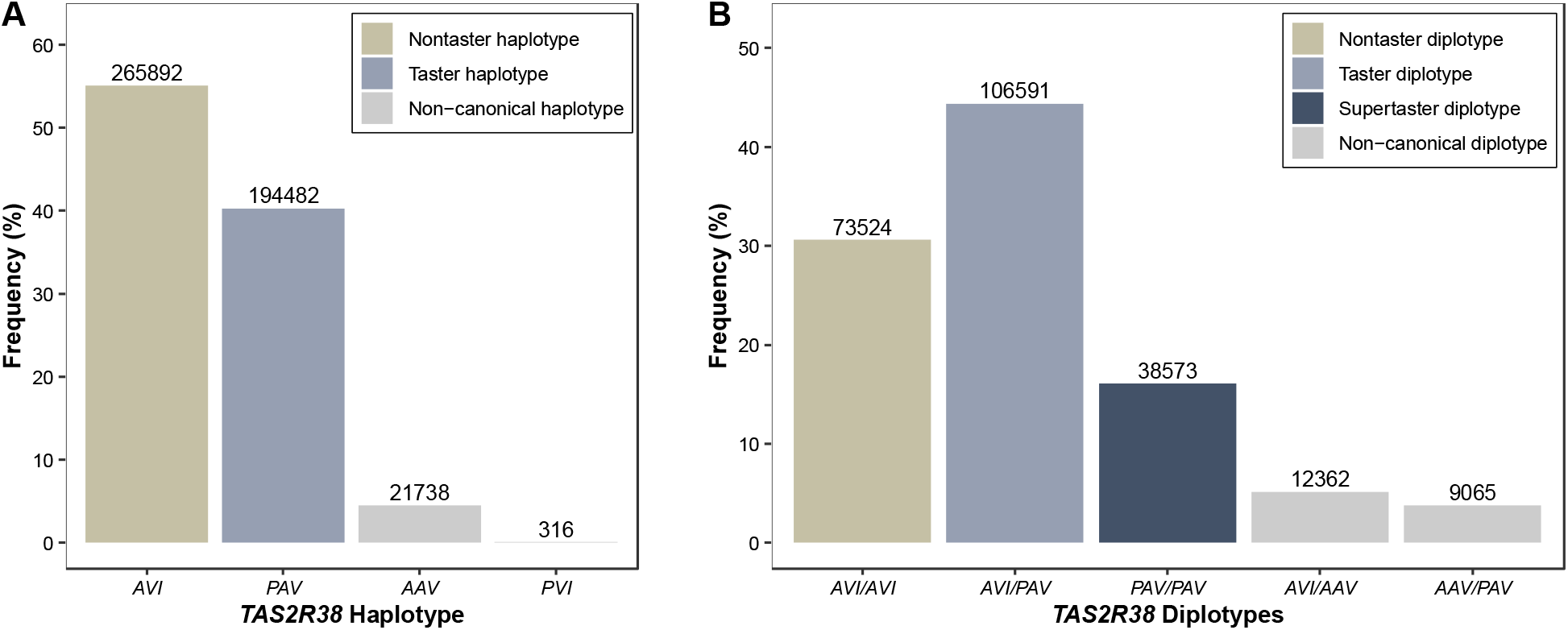
Distribution of common *TAS2R38* haplotypes (A) and diplotypes (B) among 240,115 European adults without type 2 diabetes in the UKB. Genotypes were derived from three functional variants in *TAS2R38* (rs713598, rs1726866, rs10246939); numbers above each bar reflect the *n* per genotype.

### Functional variation in the TAS2R38 bitter taste receptor

Our primary exposure was canonical *TAS2R38* diplotype based on functional variants in the receptor, as previously defined.^16,17^ Genotyping, imputation, and quality control of the UKB genetic data have been described.^25^ We selected three well-characterized functional variants in *TAS2R38*, each of which encodes for a single amino acid substitution which impacts the ligand-binding capacity (i.e., sensitivity) of the receptor: rs713598 (C>G; *p*.Ala49Pro), rs1726866 (G>A; *p*.Ala262Val), and rs10246939 (T>C; *p*.Ile296Val) (ESM Table 1).^18^ The three variants form two common haplotypes––*AVI*, a recessive haplotype encoding non-functional receptors; and *PAV*, a dominant haplotype encoding functional receptors.^16^ The two haplotypes form three common diplotypes, *AVI/AVI, AVI/PAV*, and *PAV/PAV*, where *AVI* homozygotes have blunted bitter taste perception (“nontasters”), *PAV* carriers have intermediate bitter taste perception (“tasters”), and *PAV* homozygotes have heightened bitter taste perception (“supertasters”), based on low, moderate, and high sensitivity of the bitter taste receptor to its ligands.

**Table 1.**
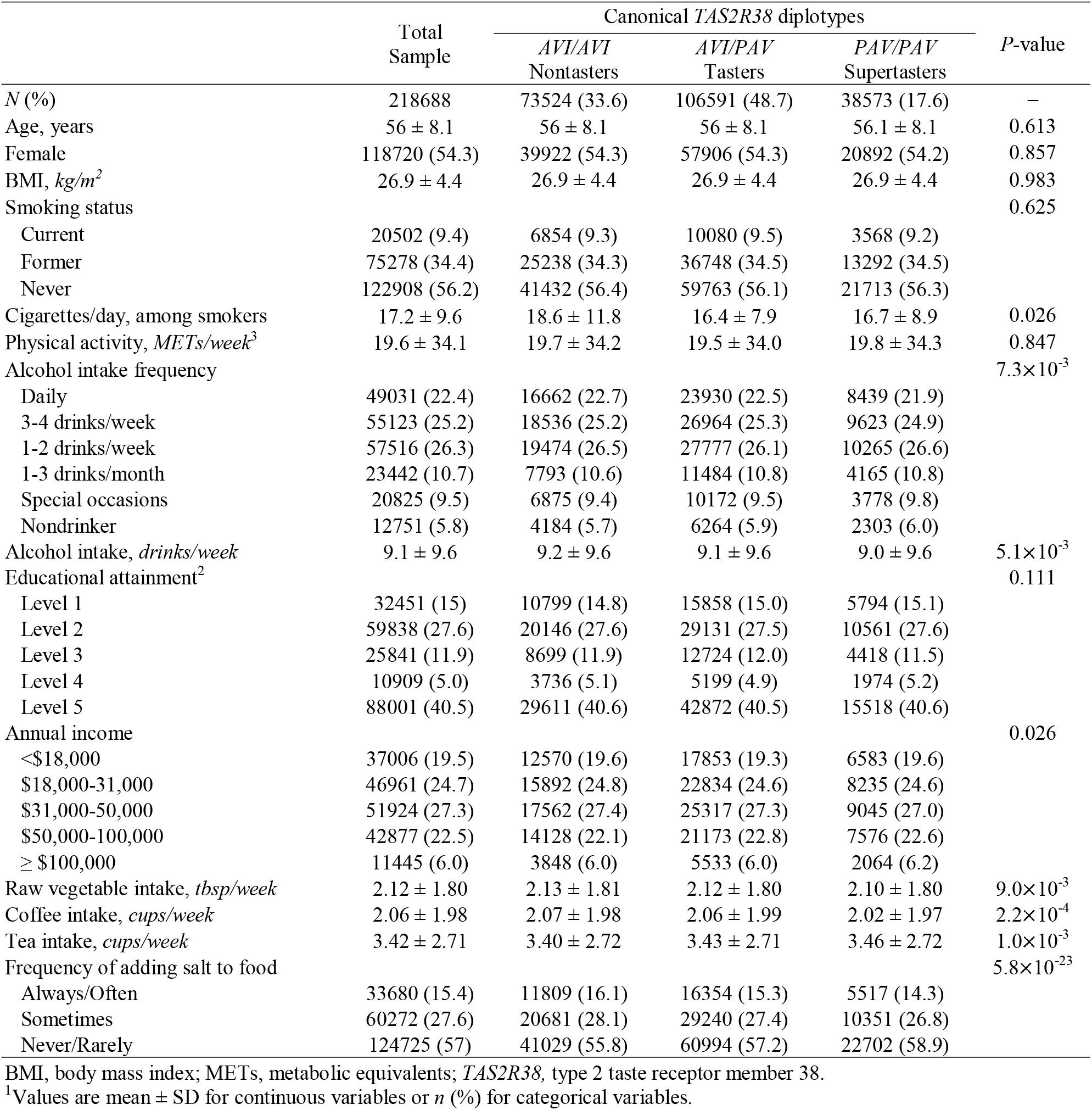
Participant characteristics across *TAS2R38* diplotypes (*N =* 218,688)^1^.

*TAS2R38* haplotypes were derived by performing haplotype phasing using SHAPEIT (*v4*.*2*)^28^ on a 2MB window surrounding the gene on chromosome 7 in the UKB European subsample. Phased haplotypes were then combined into diplotypes. To focus our analysis on functional variations with established impacts on taste receptor sensitivity, we excluded individuals with rare (<1% frequency) or non-canonical haplotypes (n = 21,427, 8.9% excluded; participant descriptions provided in ESM Table 2), and restricted the analytical sample to those with canonical *TAS2R38* diplotypes, as described above.

**Table 2.** Associations of *TAS2R38* diplotypes with random and 0-2hr glucose^1^.

|  | Random glucose (mg/dL) |  |  | 0-2hr glucose (mg/dL) |  |  |
| --- | --- | --- | --- | --- | --- | --- |
| | Beta (95% CI) | ($P_{pairwise}$)<sup>2</sup> | $P_{trend}$<sup>3</sup> | Beta (95% CI) | ($P_{pairwise}$)<sup>2</sup> | $P_{trend}$<sup>3</sup> |
| **Base model** |  |  |  |  |  | 0.003** |
| Additive *TAS2R38* diplotype, per *PAV* | -0.07 (-0.14, -0.01) | — | 0.021** | -0.24 (-0.39, -0.08) | — |  |
| AVI/AVI nontasters | Reference | — |  | Reference | — |  |
| AVI/PAV tasters | -0.05 (-0.15, 0.04) | 0.270 |  | -0.27 (-0.51, -0.03) | 0.028* |  |
| PAV/PAV supertasters | -0.15 (-0.28, -0.03) | 0.018** |  | -0.46 (-0.78, -0.14) | 0.004** |  |
| **BMI model** |  |  |  |  |  | 0.003** |
| Additive *TAS2R38* diplotype, per *PAV* | -0.07 (-0.13, -0.01) | — | 0.021** | -0.23 (-0.38, -0.08) | — |  |
| AVI/AVI nontasters | Reference | — |  | Reference | — |  |
| AVI/PAV tasters | -0.06 (-0.15, 0.04) | 0.256 |  | -0.27 (-0.50, -0.03) | 0.030* |  |
| PAV/PAV supertasters | -0.15 (-0.28, -0.02) | 0.019** |  | -0.44 (-0.76, -0.13) | 0.006** |  |

<sup>1</sup>Values are Beta (95% CI) glucose levels from generalized linear models adjusted for age, sex, 10 genetic PCs, fasting time (for random glucose, only), assessment center (Base model) and BMI (BMI model); \* $P < 0.05$ ; \*\* $P < 0.05/2$ , for 2 outcomes. The total *N* available for each outcome was 218,688 and 57,652 for random and 0-2 hr glucose, respectively.
<sup>2</sup> $P$ -value based on a categorical *TAS2R38* diplotype exposure for comparison vs. AVI/AVI nontasters (reference group).
<sup>3</sup> $P$ -value based on additive *TAS2R38* diplotype exposure for linear trends per *PAV* haplotype.

### Markers of glucose homeostasis

The primary outcomes were random glucose in mg/dL, assessed as part of routine UKB laboratory testing at baseline,^25^ measured at any time of day regardless of when participants last consumed a meal or caloric beverage; and 0-2 hr glucose in mg/dL, defined as glucose measured within 2 hours (hr) of the last consumed meal or caloric beverage based on the self-reported time since the last meal. To capture glucose variation over subsequent fasting times, we defined additional fasting windows as 3-, 4-, 5-, 6-12 and 12-24 hr (see ESM Table 3 for *n* and summary statistics for glucose levels in each interval). To minimize the impact of extreme values, we excluded participants with missing or extreme values for random glucose, considered as >5 standard deviations (SD) from the mean; and with self-reported time since last meal >24 hr (*n*=26 participants excluded; values range from 24-48 hr).

### Sociodemographic, dietary and lifestyle covariates in the UKB

Sociodemographic and lifestyle factors were measured using participant surveys at baseline.^25^ Demographic covariates included sex and age. Lifestyle covariates included smoking status, defined as current, previous, or never tobacco user; alcohol use, defined as intake frequency categories ranging from daily to “special occasions” or never; and physical activity level, defined as tertiles of excess metabolic equivalents per week.^29^ In all cases, we recoded values of “Do not know” or “Don’t want to report” as missing. For descriptive analyses, we additionally used measures of cigarettes per day (calculated among current smokers) and alcoholic drinks per day (among current drinkers).^25^ To capture socioeconomic status (SES), we used categorical variables for income level, defined by annual income brackets (range: <$18,000 to >$100,000) and educational attainment, defined as a 5-level variable based on the International Standard Classification of Education, using published methods.^30^

Dietary covariates were derived from the UKB FFQ administered at baseline using processes and procedures described previously.^31^ Briefly, the UKB FFQ assessed self-reported intake frequencies and preferences for ~20 foods, using continuous (e.g., tablespoons of vegetables) and ordinal (e.g., never, once per week, or daily servings) frequency measures as well as categorical measures of food type preferences (e.g., bread, milk, or coffee type). We converted all measures to integer variables by transforming ordinal to quantitative and categorial to binary values; and recoded responses of “Do not know” or “Prefer not to answer” as missing. Dietary variables were then median-imputed and winsorized to 5 SD, resulting in 24 continuous dietary traits for analysis.

### Statistical analysis

Consistent with prior studies, we defined *TAS2R38* diplotype as an additive, continuous exposure based on the number of *PAV* haplotypes (0 = *AVI/AVI*, 1 = *AVI/PAV*, 2 = *PAV/PAV*; primary analyses), akin to allele dosage. We also defined a categorial exposure to enable pairwise comparisons across diplotypes (*AVI/AVI* [reference] vs. *AVI/PAV* or vs. *PAV/PAV*). Descriptive statistics (mean ± SD and *n* [%]) were used to compare sociodemographic and behavioral characteristics across *TAS2R38* diplotypes. *P*-values were determined for differences in continuous and categorical variables by additive *TAS2R38* diplotype, using unadjusted linear models and Chi Square tests, respectively.

To determine the associations of *TAS2R38* diplotypes with glucose homeostasis, we used generalized linear models adjusted for age, sex, assessment center, 10 genetic PCs and fasting time, the latter of which was used to account for variability in time since the last meal as well as typical variation in glucose levels across postprandial and fasting windows (base model).^32^ All associations were also tested in a base model adjusted for BMI to account for potential confounding by adiposity (BMI-adjusted model). We summarized results as effect estimates and *P*-values on a linear scale (per *PAV* haplotype; *Ptrend*) and as estimated and SE pairwise differences in outcomes for *PAV/PAV* supertasters and *PAV/AVI* tasters vs. *AVI/AVI* nontasters (reference group; *Ppairwise*). To test for independent replication, we performed *post hoc* analyses by querying summary statistics from published GWAS of glycemic traits in European samples (excluding UKB) performed by the Meta-Analysis of Glucose and Insulin-related Traits Consortium (MAGIC). For direct comparisons with published summary statistics, we calculated effect estimates in the UKB sample at the variant-level using the BMI-adjusted model and aligned the effect alleles for both cohorts to the allele associated with higher bitter taste perception.

To test for behaviorally-independent associations, we first selected dietary and lifestyle (i.e., behavioral) covariates based on prior relations to *TAS2R38* diplotypes or glucose homeostasis. We confirmed the quality of covariates for capturing variability in blood glucose compared to covariate-only BMI-adjusted models using likelihood ratio tests. We then quantified ‘direct’ effects of TAS2R38 diplotype on glucose levels after adjusting for behavioral covariates and calculated percent change in estimates upon adjustment. Second, we used negative control tests to confirm a lack of associations with glucose homeostasis of variants in other bitter taste receptors, related to similar dietary and lifestyle behaviors as TAS2R38, but with no proposed actions in glucose metabolism. We selected variants with genome-wide significant associations to bitter taste perception (rs10772420 in *TAS2R14* related to quinine perception; rs2597979 in *TAS2R19* related to caffeine perception^33^), coded them as additive exposures based on allele dosage for higher taste perception (A for rs1077240; G for rs2597979) and tested their associations with glucose levels in BMI-adjusted models to compare with TAS2R38 variant-level estimates.

Statistical significance was defined used an *a priori* threshold of *P* <0.025 (0.05/2) to correct for multiple comparisons of two primary outcomes. All analyses were performed in R (*v*4.1) and code used to perform the analyses have been made available on GitHub (https://github.com/julieeg/taste2d).

## Results

In total, 240,115 individuals with an average age of 57 ± 14 years (median ± IQR) and BMI of 26.3 ± 5.3 kg/m^2^ had available data for analysis (Table 1, ESM Fig. 1). There were no differences in clinical and behavioral characteristics between included participants and those excluded based on rare or non-canonical diplotypes (*n* = 21,427; ESM Table 2). Among included participants, 34%, 49% and 18% were categorized as *AVI/AVI* “nontasters”, *PAV/AVI* “tasters” and *PAV/PAV* “supertasters”, respectively (Fig. 1; henceforth referred to using the descriptive label).

Across canonical *TAS2R38* diplotypes, there were no differences in age, sex, BMI, or education level (Table 1). Compared to nontasters, supertasters consumed fewer alcoholic beverages per week (*Ppairwise* = 0.006); and among current smokers, tasters and supertasters (i.e., *PAV* carriers) reported fewer cigarettes per day (*Ppairwise* = 0.019), consistent with expected associations between *TAS2R38* diplotypes and health-related behaviors.^20–23^ When assessed as an additive exposure, we further confirmed expected associations of PAV haplotypes with less alcohol and cigarette use (*Ptrend =* 7.3×10^−3^ and 0.026), less addition of salt to foods (*Ptrend* = 7.7×10^−15^), lower intake of coffee and raw vegetables (*Ptrend* = 2.7×10^−4^ and 0.008, respectively), and higher intake of tea (*Ptrend* = 7.5×10^−4^).

### Associations of TAS2R38 diplotype with glucose homeostasis

To investigate the influence of functional variants in *TAS2R38* on glucose homeostasis, we first determined the associations of *TAS2R38* diplotypes with random glucose measured at any time. We found significant, inverse associations between each additional *PAV* haplotype and glucose levels (*Ptrend* = 0.021), which corresponded to an estimated mean difference in random glucose levels of −0.15 (95% CI: −0.28, −0.02) mg/dL between supertasters and nontasters (reference) in BMI-adjusted models (*Ppairwise* = 0.019) (Table 2).

We next evaluated the associations between *TAS2R38* diplotype with 0-2hr glucose as a marker of postprandial glucose responses. Within the 0-2hr time frame, each additional *PAV* haplotype associated with a change of −0.24 (95% CI: −0.39, −0.08) mg/dL (*Ptrend* = 0.003), reflecting a greater magnitude and strength of TAS2R38 effects on 0-2 hr glucose than random glucose (beta = −0.07 mg/dL; *Ptrend* = 0.021) (Table 2). Pairwise differences for supertasters vs. nontasters were also significant below the study-wide threshold (beta = −0.46; *Ppairwise* = 0.004), while differences for tasters and supertasters reached nominal significance with intermediate effect estimates (beta = −0.27; *Ppairwise* = 0.028), both of which robust to BMI adjustment (*Ppairwise* = 0.006 and 0.03, respectively). (Table 2).

To evaluate whether these associations were specific to the 0-2 hr time window, we determined the associations between *TAS2R38* diplotype and glucose levels over subsequent time windows (3, 4, 5, 6-12 hr) (ESM Table 4). In BMI-adjusted models, we found no significant associations between *TAS2R38* diplotype and glucose levels in any later time window (beta [95% CI] per PAV haplotype combined for all times > 2hr = −0.016 [−0.08, 0.05]; *Ptrend* = 0.629). For more interpretable estimates, we calculated estimated marginal mean glucose levels for individuals with each *TAS2R38* diplotype within each fasting window (Fig 2a); and for 0-2 hr glucose, based on number of bitter taste increasing alleles for each variant (Fig 2b). These data reaffirmed that the observed associations of *TAS2R38* with glucose levels occurred specifically in the first 2 hr following a meal.

**Figure 2.**
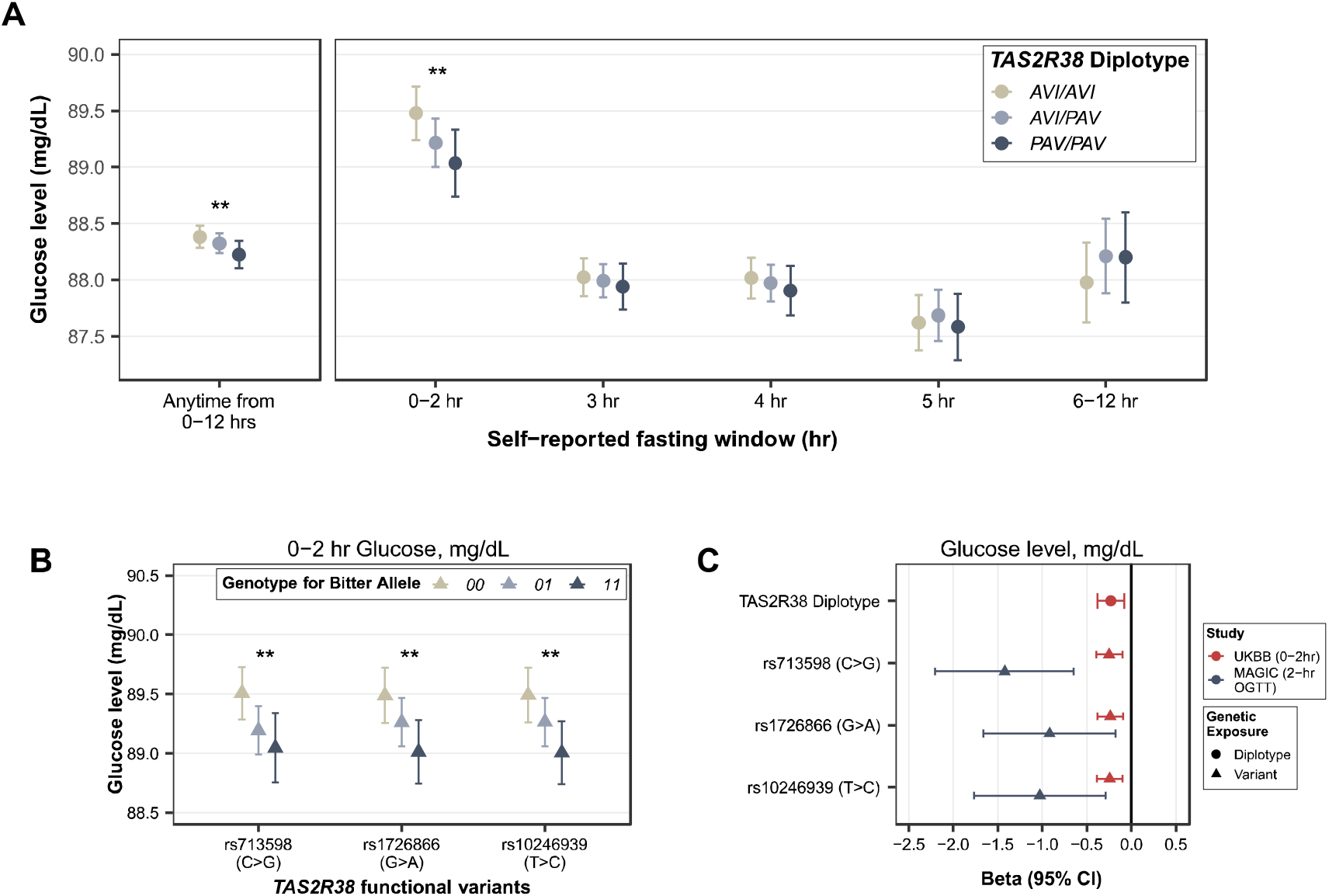
Associations of functional variants in *TAS2R38* with postprandial glycemia. Values are from linear models adjusted for age, sex, 10 genetic PCs, assessment center and BMI. Glucose levels are summarized as estimated marginal means over each time window by *TAS2R38* diplotype (**A**) and in the 0-2 hr time window by number of bitter taste increasing alleles for each functional variant (**B**). Associations of *TAS2R38* diplotype and functional variants with 0-2hr glucose (UKB) and glucose levels at 2-hr after a 75 g OGTT (MAGIC) are shown as evidence of independent replication (**C**). For panels A and B, asterisks denote statistical significance; ^**^*P* <0.025, study-wide significance threshold.

To test for independent replication, we queried summary statistics from a meta-analysis of 9 discovery GWAS for 2-hr glucose after a 75 g OGTT (gold standard postprandial glucose assessments) performed by the MAGIC consortium (*N* = 24,611 individuals of European ancestry without type 2 diabetes)^34^ to compared estimates with to those obtained at the variant-level in the UKB. All three *TAS2R38* functional variants were significantly associated with lower 2-hr OGTT glucose (Fig 2c), providing evidence of replication in an independent cohort with a direct assessment of 2-hr OGTT glucose levels.

### Assessing the potential for behaviorally-mediated effects of TAS2R38 on glucose homeostasis

As functional variants in *TAS2R38* have been implicated in dietary and lifestyle behaviors that may contribute to glucose metabolism,^35^ we assessed whether the observed associations with postprandial glucose could be “behaviorally-mediated.” As noted above, we observed expected relations of TAS2R38 diplotypes with dietary and lifestyle behaviors in our sample (Table 1; ESM 1); and also confirmed the quality of behavioral covariates for capturing variability in 0-2hr glucose compared to a covariate-only BMI-adjusted model (LRT *P*-value for diet+lifestyle model = 1.98×10^−58^; Fig 3a). We then tested the robustness of associations between *TAS2R38* and 0-2 hr glucose to sequential adjustment for behavioral covariates, finding that additive TAS2R38 associations and pairwise differences for supertasters vs. nontasters remained significant with marginal changes in effect estimates: coefficients changes by −3.4% to 1.03% after adjustment (lifestyle + diet model, beta [95% CI] per *PAV* haplotype = −0.22 [−0.38, −0.07]; *Ptrend* = 4.0×10^−3^; beta [95% CI] difference for supertasters vs. nontasters = −0.44 [−0.75, −0.12]; *Ppairwise* = 0.006) (Fig 3b).

**Figure 3.**
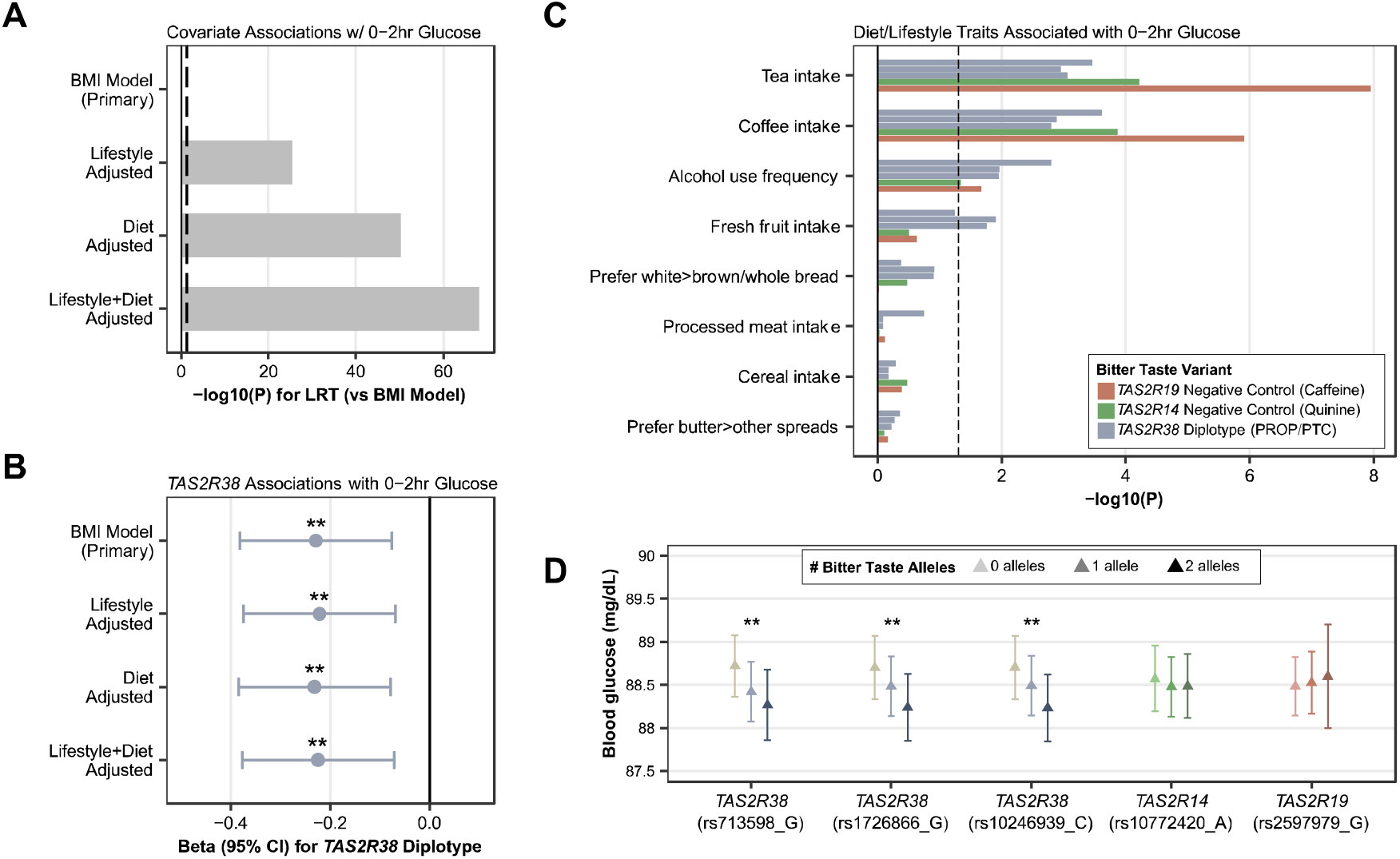
Behaviorally-independent associations of *TAS2R38* with 0-2 hr glucose. Dietary and lifestyle covariates significantly associated with 0-2 hr glucose relative to a covariate-only BMI-adjusted model (**A**), and sequentially adding behavioral covariates to BMI-adjusted models had marginal effects on the magnitude and strength of associations between *TAS2R38* and 0-2 hr glucose (**B**). Associations from BMI-adjusted models of variants in other bitter taste receptors related to caffeine and quinine perception with dietary and lifestyle behaviors relevant to 0-2 hr glucose as *TAS2R38* diplotypes (**C**). Estimated marginal mean (95% CI) blood glucose levels based on number of bitter taste increasing alleles differed significantly only for variants in the *TAS2R38* receptor, and not for negative control variants in other bitter taste receptors (**D**).

To further inform the plausibility for these effects to be behaviorally-independent, we selected variants in two other bitter taste receptor genes––known to associate with bitter taste perception, and similar dietary and lifestyle traits as TAS2R38 but without proposed functions in glucose homeostasis––to serve as negative controls: rs10772420 in *TAS2R14* and rs2597979 in *TAS2R19*. We confirmed that both variants showed similar patterns of associations as TAS2R38 variants to dietary and lifestyle traits relevant to glucose homeostasis (Fig 3c). However, we found that neither rs10772420 nor rs2597979 showed significant associations with 0-2 hr glucose in BMI-adjusted models, nor in models additionally adjusted for dietary and lifestyle behaviors (lifestyle + diet model *Ptrend* = 0.595 and 0.615, respectively; Fig. 3d).

Finally, based on prior findings that *TAS2R38* functional variants may have stronger associations with postprandial glucose levels among males than females,^23^ we ran exploratory sex-stratified analyses. We found that associations of additive and categorical *TAS2R38* diplotypes and 0-2hr glucose were only significant among males with step-wise differences per PAV haplotype (beta [95% CI] = −0.34 (−0.56, −0.11), *Ptrend* = 0.003 vs. −0.14 (−0.35, 0.07); 0.195 for females), while in females, we noted that tasters and supertasters (i.e. PAV carriers) had similarly lower postprandial glucose levels than nontasters (ESM Table 5), consistent with inverse associations between TAS2R38 sensitivity and 0-2hr glucose levels.

## Discussion

In a large-scale analysis among European adults without type 2 diabetes, we found that functional variants in TAS2R38 were significantly associated with postprandial glucose levels. Specifically, PAV/PAV supertasters had significantly lower 0-2 hr ‘postprandial’ glucose levels than AVI/AVI nontasters, with stepwise reductions in glucose levels corresponding to the number of PAV haplotypes. These associations were replicated in independent GWAS of OGTT glucose responses and remained robust after adjusting for dietary and lifestyle factors, consistent with our hypothesis and prior experimental evidence of direct TAS2R38 actions in postprandial glucose metabolism.

Our findings support emerging evidence implicating TAS2R38 receptors in glucose homeostasis. Preclinical studies show that supplementation or intragastric infusion with bitter-tasting ligands of type 2 receptors can elicit GLP-1 secretion and lower blood glucose in response to mixed-nutrient challenges in healthy individuals and those with type 2 diabetes.^12–14^ In the one prior study evaluating the associations of *TAS2R38* functional variations and glucose homeostasis, in ~1,000 German adults (60% female) without type 2 diabetes, genetic tasters and supertasters (i.e., *PAV* carriers) had significantly lower 2-hr AUC glucose than nontasters after a 75 g OGTT.^23^ Our findings in ~210,000 adults of European ancestry without type 2 diabetes were consistent with these observations. We further parsed out the effects by diplotype, showing step-wise decreases in postprandial glucose levels by PAV haplotype, reflecting a dose-response relationship based on TAS2R38 receptor sensitivity.

By using large-scale individual-level data to account for behavioral factors, we provided human study data to support behaviorally-independent actions of TAS2R38 in glucose homeostasis. As expected,^21–23^ supertasters reported lower alcohol and cigarette use and less frequent fruit and vegetable intake, which could contribute to glucose metabolism.^35,36^ While these covariates were significantly related to glucose levels in our study, adjusting for them did not attenuate the TAS2R38-glucose associations. Moreover, variants in other bitter taste receptor genes, related to bitter taste perception^33^ and similar health-related habits as TAS2R38 but without proposed roles in glucose homeostasis (i.e., negative controls), had no effect on glucose levels, limiting plausibility for health-related behaviors to explain the associations.

Based on experimental evidence in mice and human cell models,^4,7–10,10,11^ the mechanism proposed to underly the observed associations is a TAS2R38-mediated G-protein signaling cascade in enteroendocrine L-cells in the gut, initiated upon the consumption of foods or beverages, which triggers the secretion of GLP-1 and leads to a reduction in blood glucose levels. While we could not measure GLP-1 levels in our study, the dose-response relationship (based on TAS2R38 receptor sensitivity), time-specificity of associations to postprandial windows (when most GLP-1 is secreted^37^), and robustness of associations to behavioral adjustment are consistent with this as an explanatory mechanism.

Nevertheless, as GLP-1 is also stored and secreted from oral taste receptor cells,^38^ it could be speculated that greater TAS2R38 sensitivity may increase GLP-1 paracrine signals in the mouth, leading to greater satiety signals and lower food intake which would require less compensatory glycemic exertions.^39^ This may have contributed to the differences in glucose responses observed in the UKB, as participants were allowed to eat freely and did not report on the amount of foods consumed prior to glucose assessment. However, this is unlikely to explain differences in OGTT glucose responses in independent GWAS of postprandial glucose responses, for which standardized 75 g glucose challenges were provided that could not be influenced by oral satiety signals.^34^

In exploratory analyses by sex, we found preliminary evidence to support stronger associations of TAS2R38 functional variants and postprandial glycemia in males than females. This is consistent with evidence from the Sorbs cohort (~1,000 German adults; described above) which found, upon stratifying by sex, that associations of TAS2R38 diplotype with lower postprandial glucose levels were only significant among males. Here, while not statistically significant, we also observed an inverse trend in postprandial glucose levels by TAS2R38 sensitivity in females. However, the degree of differences in glucose levels compared to nontasters was similar among tasters and supertasters (i.e., PAV carriers), contrary to the stepwise decreases observed in males.

From a clinical perspective, our findings may suggest that genetic tasters and supertasters have stronger postprandial GLP-1 responses than nontasters, particularly to bitter-tasting foods or ligands. This notion fits with historical evidence ascribing glycemic benefits to bitter-tasting compounds, which are now thought to stem from bitter taste receptor actions in GLP-1 signaling.^8,40^ Berberine, for example, is a bitter-tasting compound reported to have similar hypoglycemic effects as Metformin,^40^ which has been shown to act via TAS2R38-mediated GLP-1 secretion.^7,11^ Quercetin, a natural flavonoid that may lower blood glucose, was also shown to bind gut-expressed TAS2R38 receptors and enhance GLP-1 secretion *in vitro*, though these effects were attenuated by TAS2R38 inhibition.^10^ Thus, functional variants in TAS2R38 may modulate the glycemic benefit from these compounds, providing targets to advance precision medicine. TAS2R38 agonists may also provide new treatment and prevention strategies for type 2 diabetes, in line with burgeoning interest in GLP-1 receptor agonists that augment GLP-1 signaling.^41,42^

Our results must be interpreted in the context of several limitations. First, we used a candidate gene study to examine a single genetic locus; and we identified associations did not pass a genome-wide significance threshold (*P* <5×10^−8^). However, this design was leveraged to test a hypothesis rooted in preclinical data; and we provided an independent replication in GWAS of OGTT glucose (the gold standard assessment of postprandial glucose responses).^34^ As this was an observational study, we could not infer causality; and without objective postprandial GLP-1 measurements, we could not explicitly test proposed mechanisms surrounding GLP-1 modulation. While we leveraged a negative control paradigm to strengthen our ability to infer causality, the selected variants may serve as imperfect controls as they respond to different bitter stimuli^33^ and may have yet undiscovered roles in glucose metabolism or nutrient sensing in other peripheral tissues.^3,5,6^ While we aimed to robustly adjust for dietary and lifestyle behaviors, the covariates were derived from brief FFQs or self-report surveys which are subject to recall bias and confounding. To minimize the risk of measurement error we performed a complete case analysis, although this might have introduced sampling bias. Finally, our focus on individuals of European ancestry limits our ability to generalize findings to other ancestry groups, among whom the frequency of *TAS2R38* genotypes, dietary habits, and their phenotypic consequences might differ.^43,44^

## Conclusions

In a large cohort of adults without type 2 diabetes, functional variants encoding more sensitive TAS2R38 bitter taste receptors associate with lower postprandial glucose levels, independent of lifestyle or dietary behaviors. Future studies are needed to examine the direct influence of TAS2R38 functional variants on GLP-1 secretion and postprandial glucose responses to standardized and mixed meals to define the magnitude of this effect and inform on the potential for TAS2R38 to serve as a therapeutic target in precision prevention and treatment of type 2 diabetes.

## Supporting information

Electronic Supplementary Material (ESM)

## Data Availability

No new genetic or phenotypic data have been generated for this study. The UK Biobank data, including genetic and phenotypic data, are under controlled access but can be obtained through application at https://www.ukbiobank.ac.uk/. UK Biobank will consider data applications from bona fide researchers for health-related research that is in the public interest.

## Code Availability

Code supporting the conclusion of this manuscript can be found at: https://github.com/julieeg/taste2d.

## Abbreviations

AUC: area under the curve
FFQ: Food frequency questionnaire
GLP-1: Glucagon-like peptide-1
GWAS: Genome-wide association studies
OGTT: Oral glucose tolerance tests
TAS2R38: Taste receptor type 2 member 38
UKB: UK Biobank

## References

1. Drucker, D. J. GLP-1-based therapies for diabetes, obesity and beyond. Nat. Rev. Drug Discov. 1–20 (2025) doi:10.1038/s41573-025-01183-8.

2. Holst, J. J., Gribble, F., Horowitz, M. & Rayner, C. K. Roles of the Gut in Glucose Homeostasis. Diabetes Care 39, 884–892 (2016).

3. Calvo, S. S.-C. & Egan, J. M. The endocrinology of taste receptors. Nat. Rev. Endocrinol. 11, 213–227 (2015).

4. Wu, S. V. et al. Expression of bitter taste receptors of the T2R family in the gastrointestinal tract and enteroendocrine STC-1 cells. Proc. Natl. Acad. Sci. 99, 2392–2397 (2002).

5. Sternini, C. & Rozengurt, E. Bitter taste receptors as sensors of gut luminal contents. Nat. Rev. Gastroenterol. Hepatol. 22, 39–53 (2025).

6. Egan, J. M. Physiological Integration of Taste and Metabolism. N. Engl. J. Med. 390, 1699–1710 (2024).

7. Yu, Y. et al. Berberine induces GLP-1 secretion through activation of bitter taste receptor pathways. Biochem. Pharmacol. 97, 173–177 (2015).

8. Pham, H. et al. A bitter pill for type 2 diabetes? The activation of bitter taste receptor TAS2R38 can stimulate GLP-1 release from enteroendocrine L-cells. Biochem. Biophys. Res. Commun. 475, 295–300 (2016).

9. Jang, H.-J. et al. Gut-expressed gustducin and taste receptors regulate secretion of glucagon-like peptide-1. Proc. Natl. Acad. Sci. 104, 15069–15074 (2007).

10. Wang, C. et al. Quercetin Enhances GLP-1 Secretion via TAS2R38-Mediated PLC Signaling in Enteroendocrine L-Cells. Mol. Nutr. Food Res. n/a, e70109.

11. Yue, X., Liang, J., Gu, F., Du, D. & Chen, F. Berberine activates bitter taste responses of enteroendocrine STC-1 cells. Mol. Cell. Biochem. 447, 21–32 (2018).

12. Mohammadpour, Z. et al. The effect of post-oral bitter compound interventions on the postprandial glycemia response: A systematic review and meta-analysis of randomised controlled trials. Clin. Nutr. 43, 31–45 (2024).

13. Bitarafan, V. et al. Intragastric administration of the bitter tastant quinine lowers the glycemic response to a nutrient drink without slowing gastric emptying in healthy men. Am. J. Physiol.-Regul. Integr. Comp. Physiol. 318, R263–R273 (2020).

14. Bitarafan, V. et al. Dose-related effects of intraduodenal quinine on plasma glucose, glucoregulatory hormones and gastric emptying of a nutrient drink, and energy intake, in men with type 2 diabetes: a double-blind, randomised, crossover study. Diabetologia (2024) doi:10.1007/s00125-024-06344-9.

15. Rezaie, P., Bitarafan, V., Horowitz, M. & Feinle-Bisset, C. Effects of Bitter Substances on GI Function, Energy Intake and Glycaemia-Do Preclinical Findings Translate to Outcomes in Humans? Nutrients 13, 1317 (2021).

16. Kim, U. et al. Positional Cloning of the Human Quantitative Trait Locus Underlying Taste Sensitivity to Phenylthiocarbamide. Science 299, 1221–1225 (2003).

17. Mennella, J. A., Pepino, M. Y., Duke, F. F. & Reed, D. R. Psychophysical Dissection of Genotype Effects on Human Bitter Perception. Chem. Senses 36, 161–167 (2011).

18. Tan, J., Abrol, R., Trzaskowski, B. & Goddard, W. A. I. 3D Structure Prediction of TAS2R38 Bitter Receptors Bound to Agonists Phenylthiocarbamide (PTC) and 6-n-Propylthiouracil (PROP). J. Chem. Inf. Model. 52, 1875–1885 (2012).

19. Knaapila, A. et al. Genetic Analysis of Chemosensory Traits in Human Twins. Chem. Senses 37, 869–881 (2012).

20. Cornelis, M. C. & van Dam, R. M. Genetic determinants of liking and intake of coffee and other bitter foods and beverages. Sci. Rep. 11, 23845 (2021).

21. Duffy, V. B. et al. Bitter Receptor Gene (TAS2R38), 6-n-Propylthiouracil (PROP) Bitterness and Alcohol Intake. Alcohol. Clin. Exp. Res. 28, 1629–1637 (2004).

22. Calancie, L. et al. TAS2R38 Predisposition to Bitter Taste Associated with Differential Changes in Vegetable Intake in Response to a Community-Based Dietary Intervention. G3 GenesGenomesGenetics 8, 2107–2119 (2018).

23. Keller, M. et al. TAS2R38 and Its Influence on Smoking Behavior and Glucose Homeostasis in the German Sorbs. PLOS ONE 8, e80512 (2013).

24. Sudlow, C. et al. UK biobank: an open access resource for identifying the causes of a wide range of complex diseases of middle and old age. PLoS Med. 12, e1001779 (2015).

25. Bycroft, C. et al. The UK Biobank resource with deep phenotyping and genomic data. Nature 562, 203–209 (2018).

26. Karczewski, K. J. et al. Pan-UK Biobank GWAS improves discovery, analysis of genetic architecture, and resolution into ancestry-enriched effects. 2024.03.13.24303864 Preprint at 10.1101/2024.03.13.24303864 (2024).

27. Eastwood, S. V. et al. Algorithms for the Capture and Adjudication of Prevalent and Incident Diabetes in UK Biobank. PLOS ONE 11, e0162388 (2016).

28. Delaneau, O., Zagury, J.-F., Robinson, M. R., Marchini, J. L. & Dermitzakis, E. T. Accurate, scalable and integrative haplotype estimation. Nat. Commun. 10, 5436 (2019).

29. Guo, W., Bradbury, K. E., Reeves, G. K. & Key, T. J. Physical activity in relation to body size and composition in women in UK Biobank. Ann. Epidemiol. 25, 406–413.e6 (2015).

30. Ge, T. et al. The Shared Genetic Basis of Educational Attainment and Cerebral Cortical Morphology. Cereb. Cortex 29, 3471–3481 (2019).

31. Westerman, K. E. et al. Genome-wide gene–diet interaction analysis in the UK Biobank identifies novel effects on hemoglobin A1c. Hum. Mol. Genet. 30, 1773–1783 (2021).

32. Lagou, V. et al. GWAS of random glucose in 476,326 individuals provide insights into diabetes pathophysiology, complications and treatment stratification. Nat. Genet. 55, 1448–1461 (2023).

33. Hwang, L.-D. et al. Bivariate genome-wide association analysis strengthens the role of bitter receptor clusters on chromosomes 7 and 12 in human bitter taste. BMC Genomics 19, 678 (2018).

34. Saxena, R. et al. Genetic variation in GIPR influences the glucose and insulin responses to an oral glucose challenge. Nat. Genet. 42, 142–148 (2010).

35. Berry, S. E. et al. Human postprandial responses to food and potential for precision nutrition. Nat. Med. 26, 964–973 (2020).

36. Huber, H., Schieren, A., Holst, J. J. & Simon, M.-C. Dietary impact on fasting and stimulated GLP-1 secretion in different metabolic conditions – a narrative review. Am. J. Clin. Nutr. 119, 599–627 (2024).

37. Xie, C. et al. Plasma GLP-1 Response to Oral and Intraduodenal Nutrients in Health and Type 2 Diabetes—Impact on Gastric Emptying. J. Clin. Endocrinol. Metab. 107, e1643–e1652 (2022).

38. Shin, Y.-K. et al. Modulation of taste sensitivity by GLP-1 signaling. J. Neurochem. 106, 455–463 (2008).

39. Martin, B. et al. Modulation of taste sensitivity by GLP-1 signaling in taste buds. Ann. N. Y. Acad. Sci. 1170, 98–101 (2009).

40. Chou, W.-L. Therapeutic potential of targeting intestinal bitter taste receptors in diabetes associated with dyslipidemia. Pharmacol. Res. 170, 105693 (2021).

41. Gratzl, S., Rodriguez, P. J., Cartwright, B. M. G., Baker, C. & Stucky, N. L. Monitoring Report: GLP-1 RA Prescribing Trends - December 2023 Data. http://medrxiv.org/lookup/doi/10.1101/2024.01.18.24301500 (2024) doi:10.1101/2024.01.18.24301500.

42. Watanabe, J. H., Kwon, J., Nan, B. & Reikes, A. Trends in glucagon-like peptide 1 receptor agonist use, 2014 to 2022. J. Am. Pharm. Assoc. 0, (2023).

43. Risso, D. S. et al. Global diversity in the TAS2R38 bitter taste receptor: revisiting a classic evolutionary PROPosal. Sci. Rep. 6, 25506 (2016).

44. Chupeerach, C. et al. The influence of TAS2R38 bitter taste gene polymorphisms on obesity risk in three racially diverse groups. BioMedicine 11, 43–49 (2021).

