## Supplementary material for "Functional variants in the TAS2R38 bitter taste receptor associate with postprandial glycemia": Electronic Supplementary Material (ESM)

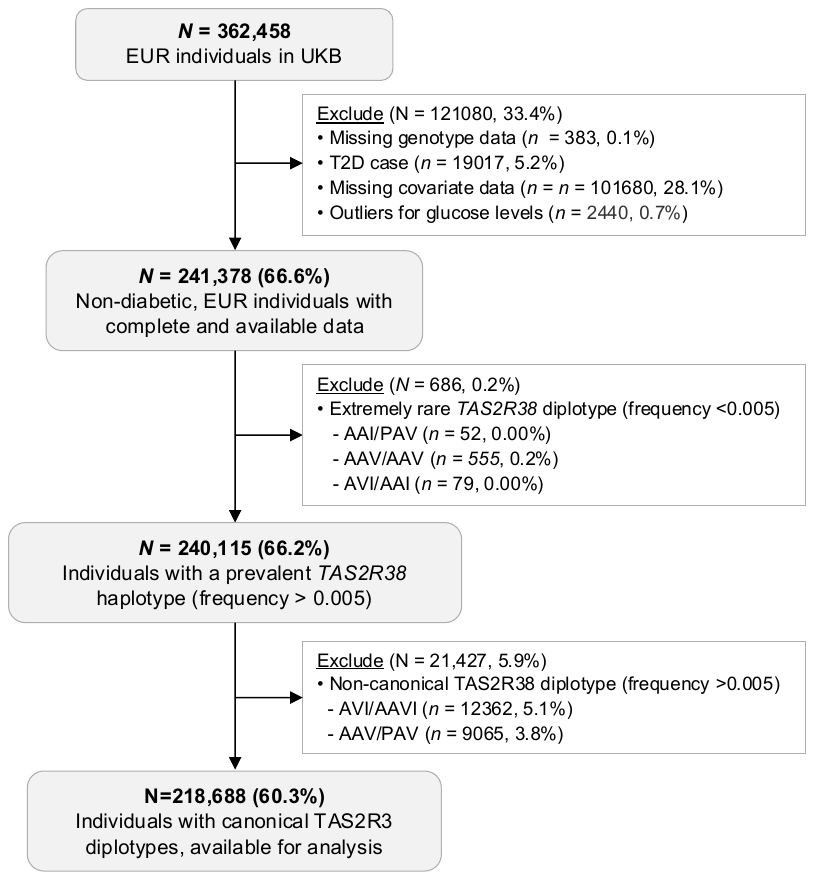

**ESM Figure 1**. Flow chart of included participants from the UK Biobank

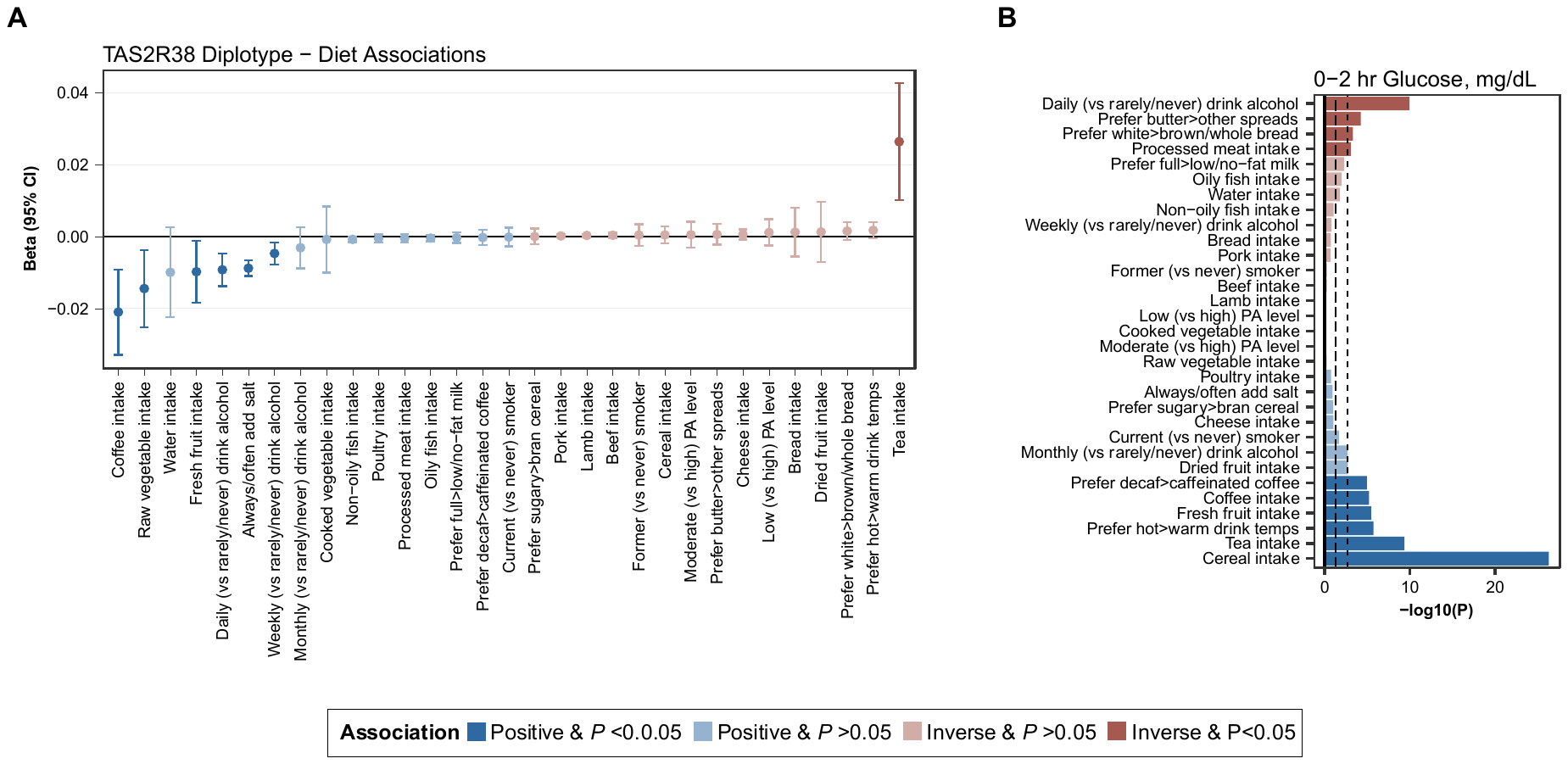

**ESM Figure 2.** Associations of lifestyle and dietary covariates with TAS2R38 diplotype and 0-2hr glucose (mg/dL). Estimates were generated from generalized linear models adjusting for age, sex, 10 genetic PCs, and BMI (kg/m^2^) using additive TAS2R38 diplotypes as a continuous exposure with each covariate outcome (A) or using each covariates as an independent exposure with 0-2 hr glucose (mg/dL) as a continuous outcome (B).

| **ESM Table 1.** Functional variants in the *TAS2R38* bitter taste receptor gene comprising canonical TAS2R38 haplotypes and diplotypes^1^ | | | | | | |
| --- | --- | --- | --- | --- | --- | --- |
| Variant | Chr:Position^1^ | Ref>Alt | Alt AF | Coding Consequence | **Common Haplotypes** | |
|  |  |  |  |  | *AVI* | *PAV* |
| rs713598 | 7:141973545 | C>G | 0.403 | p.Ala49Pro | C (Alanine) | G (Proline) |
| rs1726866 | 7:141972905 | G>A | 0.554 | p.Ala262Val | A (Valine) | G (Alanine) |
| rs10246939 | 7:141972804 | T>C | 0.446 | p.Ile296Val | T (Isoleucine) | C (Valine) |
| ^1^Chromosome and position are reported for GRCh38; AF reflects allele frequency for the alternative allele in the UKB European subsample. | | | | | | |

| **ESM Table 2.** Participant characteristics across canonical and rare TAS2R38 diplotypes (*N =* 240,115)^1^ | | | | | | |
| --- | --- | --- | --- | --- | --- | --- |
|  | Canonical Diplotypes | | |  | Other Diplotypes | |
|  | *AVI/AVI* | *AVI/PAV* | *PAV/PAV* |  | *AVI/AAV* | *AAV/PAV* |
| *N* (%) | 73524 (31%) | 106591 (44%) | 38573 (16%) |  | 12362 (5%) | 9065 (4%) |
| Age, years | 56 ± 8.1 | 56 ± 8.1 | 56.1 ± 8.1 |  | 56.1 ± 8.1 | 56.1 ± 8.1 |
| Female | 39922 (54%) | 57906 (54%) | 20892 (54%) |  | 6691 (54%) | 4899 (54%) |
| BMI, kg/m^2^ | 26.9 ± 4.4 | 26.9 ± 4.4 | 26.9 ± 4.4 |  | 26.9 ± 4.4 | 26.9 ± 4.4 |
| Smoking status |  |  |  |  |  |  |
| Current | 6854 (9%) | 10080 (9%) | 3568 (9%) |  | 1184 (10%) | 885 (10%) |
| Former | 25238 (34%) | 36748 (34%) | 13292 (34%) |  | 4264 (34%) | 3030 (33%) |
| Never | 41432 (56%) | 59763 (56%) | 21713 (56%) |  | 6914 (56%) | 5150 (57%) |
| Cigarettes/day, among smokers | 18.6 ± 11.8 | 16.4 ± 7.9 | 16.7 ± 8.9 |  | 18.5 ± 9.9 | 13.8 ± 6.1 |
| Physical activity level, MET/wk | 33.8 ± 43.3 | 33.7 ± 43.1 | 33.7 ± 43.2 |  | 34 ± 43.6 | 33.8 ± 42.9 |
| Alcohol intake frequency |  |  |  |  |  |  |
| Daily | 16662 (23%) | 23930 (22%) | 8439 (22%) |  | 2820 (23%) | 1945 (21%) |
| 1-4 drinks/week | 38010 (52%) | 54741 (51 %) | 19888 (52%) |  | 6437 (52%) | 4756 (52%) |
| 1-3 drinks/month | 7793 (11%) | 11484 (11%) | 4165 (11%) |  | 1269 (10%) | 1008 (11%) |
| Special occasions | 6875 (9%) | 10172 (10%) | 3778 (10%) |  | 1134 (9%) | 848 (9%) |
| Nondrinker | 4184 (6%) | 6264 (6%) | 2303 (6%) |  | 702 (6%) | 508 (6%) |
| Alcohol intake, *drinks/week* | 9.2 ± 9.6 | 9.1 ± 9.6 | 9 ± 9.6 |  | 9.3 ± 9.8 | 9.1 ± 9.7 |
| Education level |  |  |  |  |  |  |
| Level 1 | 29611 (40%) | 42872 (40%) | 15518 (40%) |  | 5036 (41%) | 3545 (39%) |
| Level 2 | 10799 (15%) | 15858 (15%) | 5794 (15%) |  | 1892 (15%) | 1411 (16%) |
| Level 3 | 20146 (27%) | 29131 (27%) | 10561 (27%) |  | 3255 (26%) | 2489 (27%) |
| Level 4 | 8699 (12%) | 12724 (12%) | 4418 (11%) |  | 1466 (12%) | 1097 (12%) |
| Level 5 | 3736 (5%) | 5199 (5%) | 1974 (5%) |  | 613 (5%) | 450 (5%) |
| Annual income |  |  |  |  |  |  |
| <$18,000 | 12570 (17%) | 17853 (17%) | 6583 (17%) |  | 2031 (16%) | 1567 (17%) |
| $18,000-31,000 | 15892 (22%) | 22834 (21%) | 8235 (21%) |  | 2720 (22%) | 1928 (21%) |
| $31,000-50,000 | 17562 (24%) | 25317 (24%) | 9045 (23%) |  | 2936 (24%) | 2143 (24%) |
| $50,000-100,000 | 14128 (19%) | 21173 (20%) | 7576 (20%) |  | 2411 (20%) | 1769 (20%) |
| ≥ $100,000 | 3848 (5%) | 5533 (5%) | 2064 (5%) |  | 657 (5%) | 482 (5%) |
| Raw vegetable intake, *tbsp/week* | 2.1 ± 1.8 | 2.1 ± 1.8 | 2.1 ± 1.8 |  | 2.1 ± 1.8 | 2.1 ± 1.7 |
| Coffee intake, *cups/week* | 2.1 ± 2 | 2.1 ± 2 | 2 ± 2 |  | 2.1 ± 2 | 2.1 ± 2 |
| Tea intake, *cups/week* | 3.4 ± 2.7 | 3.4 ± 2.7 | 3.5 ± 2.7 |  | 3.4 ± 2.7 | 3.5 ± 2.8 |
| Frequency of adding salt to food |  |  |  |  |  |  |
| Always/Often | 11809 (16.1%) | 16354 (15.3%) | 5517 (14.3%) |  | 2000 (16.2%) | 1348 (14.9%) |
| Sometimes | 20681 (28.1%) | 29240 (27.4%) | 10351 (26.8%) |  | 3438 (27.8%) | 2422 (26.7%) |
| Never/Rarely | 41029 (55.8%) | 60994 (57.2%) | 22702 (58.9%) |  | 6924 (56%) | 5295 (58.4%) |
| MET, metabolic equivalents per week; *TAS2R38,* taste receptor type 2 member 38.  ^1^Values are *n* (%) or mean ± SD. | | | | | | |

| **ESM Table 3.** Summary of glucose levels by self-reported time the since last meal in the UKB^1^ | | | | | | |
| --- | --- | --- | --- | --- | --- | --- |
| Fasting Category | Fasting Time (hours) | Glucose levels (mg/dL) | *n* (%) | % breakdown by TAS2R38 diplotype | | |
|  |  |  |  | AVI/AVI | AVI/PAV | PAV/PAV |
| 0-2 hr |  | 89.2 ± 13.4 | 57652 (26.4%) | (33.8%) | (48.8%) | (17.4%) |
|  | 0 | 97.1 ± 15.0 | 189 (0.09%) | (38.1%) | (45.5%) | (16.4%) |
|  | 1 | 91.8 ± 15.4 | 10148 (4.6%) | (33.3%) | (49.1%) | (17.5%) |
|  | 2 | 88.7 ± 12.8 | 47315 (21.6%) | (33.9%) | (48.7%) | (17.4%) |
| 3 hr | 3 | 88.2 ± 10.1 | 64729 (29.6%) | (33.7%) | (48.5%) | (17.9%) |
| 4 hr | 4 | 88.3 ± 8.8 | 47335 (21.6%) | (33.5%) | (48.8%) | (17.7%) |
| 5 hr | 5 | 88.1 ± 8.4 | 25831 (11.8%) | (33.8%) | (48.8%) | (17.4%) |
| 6-12hr |  | 88.2 ± 8.3 | 16977 (7.8%) | (32.9%) | (49.3%) | (17.8%) |
|  | 6 | 88.1 ± 8.2 | 11759 (5.4%) | (33.1%) | (49.1%) | (17.7%) |
|  | 7 | 88.2 ± 8.4 | 3066 (1.4%) | (31.9%) | (50.3%) | (17.8%) |
|  | 8 | 87.8 ± 8.4 | 1082 (0.5%) | (34.8%) | (48.5%) | (16.7%) |
|  | 9 | 88.3 ± 8.9 | 381 (0.2%) | (33.6%) | (47.0%) | (19.4%) |
|  | 10 | 88.9 ± 8.6 | 359 (0.2%) | (32.3%) | (51.3%) | (16.4%) |
|  | 11 | 88.9 ± 9.4 | 330 (0.2%) | (28.2%) | (51.5%) | (20.3%) |
| ≥12 hr |  | 90.0 ± 9.1 | 6164 (2.8%) | (33.5%) | (49.4%) | (17.1%) |
|  | 12 | 91.0 ± 9.0 | 1695 (0.8%) | (32.0%) | (49.2%) | (18.8%) |
|  | 13 | 91.8 ± 9.4 | 713 (0.3%) | (31.6%) | (51.9%) | (16.5%) |
|  | 14 | 90.6 ± 8.7 | 1023 (0.5%) | (34.3%) | (49.4%) | (16.3%) |
|  | 15 | 90.2 ± 9.1 | 885 (0.4%) | (34.7%) | (48.1%) | (17.2%) |
|  | 16 | 88.5 ± 8.5 | 638 (0.3%) | (32.1%) | (50.2%) | (17.7%) |
|  | 17 | 89.1 ± 9.6 | 412 (0.2%) | (35.2%) | (50.7%) | (14.1%) |
|  | 18 | 87.4 ± 8.6 | 387 (0.2%) | (36.4%) | (48.1%) | (15.5%) |
|  | 19 | 88.3 ± 8.6 | 136 (0.06%) | (39.0%) | (44.1%) | (16.9%) |
|  | 20 | 88.7 ± 9.7 | 111 (0.05%) | (35.1%) | (48.6%) | (16.2%) |
|  | 21 | 85.9 ± 8.5 | 56 (0.03%) | (28.6%) | (48.2%) | (23.2%) |
|  | 22 | 84.6 ± 9.5 | 35 (0.02%) | (37.1%) | (54.3%) | (8.6%) |
|  | 23 | 85.3 ± 8.4 | 24 (0.01%) | (12.5%) | (66.7%) | (20.8%) |
|  | 24 | 85.5 ± 9.4 | 49 (0.02%) | (44.9%) | (40.8%) | (14.3%) |
| ^1^Values are mean ± SD or *n* (%,) unless otherwise stated. | | | | | | |

| **ESM Table 4**. Associations of TAS2R38 diplotype with glucose levels (mg/dL) over ≥3hr fasting windows | | | | |
| --- | --- | --- | --- | --- |
|  | *N* | Glucose (mg/dL) | | |
|  |  | Beta (95% CI) | *P_pairwise_*^2^ | *P*_trend_^3^ |
| **3-hr Glucose** |  |  |  |  |
| Additive *TAS2R38* diplotype, per *PAV* | 64729 | -0.04 (-0.149, 0.069) | – | 0.472 |
| *AVI/AVI* nontasters | 21794 | *Reference* | – |  |
| *AVI/PAV* tasters | 31370 | -0.032 (-0.203, 0.139) | 0.710 |  |
| *PAV/PAV* supertasters | 11565 | -0.083 (-0.306, 0.14) | 0.468 |  |
| **4-hr Glucose** |  |  |  |  |
| Additive *TAS2R38* diplotype, per *PAV* | 47335 | -0.054 (-0.166, 0.057) | – | 0.339 |
| *AVI/AVI* nontasters | 15852 | *Reference* | – |  |
| *AVI/PAV* tasters | 23091 | -0.044 (-0.218, 0.131) | 0.468 |  |
| *PAV/PAV* supertasters | 8392 | -0.113 (-0.341, 0.116) | 0.333 |  |
| **5-hr Glucose** |  |  |  |  |
| Additive *TAS2R38* diplotype, per *PAV* | 25831 | -0.005 (-0.148, 0.138) | – | 0.944 |
| *AVI/AVI* nontasters | 8740 | *Reference* | – |  |
| *AVI/PAV* tasters | 12597 | 0.064 (-0.159, 0.288) | 0.571 |  |
| *PAV/PAV* supertasters | 4494 | -0.038 (-0.332, 0.257) | 0.801 |  |
| **≥6 hr Glucose** |  |  |  |  |
| Additive *TAS2R38* diplotype, per *PAV* | 23141 | 0.116 (-0.04, 0.271) | – | 0.145 |
| *AVI/AVI* nontasters | 7649 | *Reference* | – |  |
| *AVI/PAV* tasters | 11423 | 0.277 (0.034, 0.52) | 0.026^*^ |  |
| *PAV/PAV* supertasters | 4069 | 0.171 (-0.148, 0.49) | 0.294 |  |
| ^1^Values are Beta (95% CI) from generalized linear models adjusting for age, sex, 10 genetic PCs, assessment center and BMI (kg/m^2^), stratified by fasting time window.  ^2^*P*-value for pairwise comparison among categorical TAS2R38 diplotypes, relative to *AVI/AVI* nontasters (reference)  ^3^*P*-value for linear trend across additive TAS2R38 diplotypes (per PAV haplotype)  ^*^*P* <0.05; ^**^*P* <0.025 (0.05/2 outcomes) | | | | |

| **ESM Table 5.** Sex-stratified associations of TAS2R38 diplotypes with 0-2hr glucose in the BMI-adjusted model^1^ | | | | | | | | | |
| --- | --- | --- | --- | --- | --- | --- | --- | --- | --- |
|  | **Females** | | | |  | **Males** | | | |
|  | *n* | Beta (95% CI) | *P_pairwise_* | *P_trend_* |  | *n* | Beta (95% CI) | *P_pairwise_* | *P_trend_* |
| **0-2hr Glucose** |  |  |  |  |  |  |  |  |  |
| Additive *TAS2R38* diplotype, per *PAV* | 31296 | -0.14 (-0.35, 0.07) | – | 0.197 |  | 26365 | -0.34 (-0.56, -0.11) | – | 0.003^**^ |
| *AVI/AVI* nontasters | 10645 | *Reference* | – | 0.270 |  | 8844 | *Reference* | – | 0.017^**^ |
| *AVI/PAV* tasters | 15229 | -0.24 (-0.57, 0.09) | 0.151 | – |  | 12881 | -0.29 (-0.64, 0.06) | 0.100 | – |
| *PAV/PAV* supertasters | 5422 | -0.24 (-0.67, 0.20) | 0.273 | – |  | 4631 | -0.69 (-1.15, -0.23) | 0.003^**^ | – |
| *PAV* carriers vs. *AVI*/*AVI* |  |  |  |  |  |  |  |  |  |
| *AVI* homozygous (*AVI/AVI*) | 10645 | *Reference* | – | 0.116 |  | 11616 | *Reference* | – | 0.02^**^ |
| *PAV* carrier (*AVI/PAV* or *PAV/PAV*) | 20651 | -0.24 (-0.55, 0.07) | 0.128 | – |  | 17512 | 0.398 (-0.73, -0.07) | 0.018^**^ | – |
| ^1^Values are Beta (95% CI) from generalized linear model, adjusting for age, 10 genetic PCs, and BMI (kg/m^2^), stratified by sex.  ^*^*P* <0.05; ^**^*P* <0.025 (0.05/2 outcomes) | | | | | | | | | |
